# Will a natural collective immunity of Ukrainians restrain new COVID-19 waves?

**DOI:** 10.1101/2021.07.20.21260840

**Authors:** Igor Nesteruk

## Abstract

The visible and real sizes the COVID-19 epidemic in Ukraine were estimated with the use of the number of laboratory-confirmed cases (accumulated in May and June 2021), the generalized SIR-model and the parameter identification procedure taking into account the difference between registered and real number of cases. The calculated optimal value of the visibility coefficient shows that most Ukrainians have already been infected with the coronavirus, and some more than once, i.e., Ukrainians have probably achieved a natural collective immunity. Nevertheless, a large number of new strains and short-lived antibodies can cause new pandemic waves. In particular, the beginning of such a wave, we probably see in Ukraine in mid-July 2021. The further dynamics of the epidemic and its comparison with the results of mathematical modeling will be able to answer many important questions about the natural immunity and effectiveness of vaccines.

## Introduction

The early stages of the COVID-19 pandemic outbreak in Ukraine were discussed in [1, 2]. Further pandemic dynamics in this country was investigated with the use of the classical SIR model [3-5] and the statistics-based method of its parameter identification proposed in [6]. The results presented in [7-10] showed that this approach is able to predict only the first epidemic waves and only in the cases when the number of registered cases reflects the real one (fist predictions based on the initial datasets were two optimistic). Nevertheless, fairly accurate predictions about the first epidemic waves have been done for Italy, Austria, Germany, France, Spain, China, and South Korea [11].

The weakening of quarantine restrictions, changes in the social behavior and the coronavirus mutations caused changes in the epidemic dynamics and corresponding parameters of models. To detect these new epidemic waves, a simple method of numerical differentiations of the smoothed number of cases was proposed in [12, 13]. Predictions based on [3-6] for Ukraine, USA, UK, and the Republic of Moldova were not very reliable, since the second waves in these countries started before the finishing the first one [11]. To simulate different epidemic waves, the generalized SIR-model [14] and corresponding parameter identification procedure [15] were proposed. In particular, nine epidemic waves in Ukraine were calculated [11, 16, 17].

Due to the large number of asymptomatic COVID-19 patients, the actual number of infected far exceeds the number of laboratory-confirmed cases, [18-23]. In order to assess the extent of data incompleteness, the identification algorithm for SIR-parameters was modified in [24, 25]. It allowed us to determine the true sizes of the COVID-19 epidemic in Ukraine [24-26] and in Qatar [27, 28]. In this paper we present the results of SIR simulations of the new pandemic wave in Ukraine based on the dataset for the number of cases registered in the period May 25 – June 7, 2021. The visible and real dimensions of the 11-th pandemic wave in Ukraine will be estimated and discussed.

## Data

We will use the data set regarding the accumulated numbers of laboratory-confirmed COVID-19 cases in Ukraine from national sources [29, 30]. The corresponding numbers *V*_*j*_ and moments of time *t*_*j*_ (measured in days) are shown in Table 1. The values for the period *T*_*c10*_ : March 11-24, 2021 have been used in [24, 25] for SIR simulations of the tenth epidemic wave in Ukraine. Here we use the fresher dataset, corresponding to the period *T*_*c11*_: May 23 – June 5, 2021 to simulate the 11-th wave. Other *V*_*j*_ and *t*_*j*_ values will be used to control the accuracy of predictions and pandemic dynamics.

**Table 1.** Cumulative numbers of laboratory-confirmed Covid-19 cases in Ukraine *V*_*j*_ in the spring and summer of 2021 according to the national statistics, [29, 30].

| Day in corresponding month of 2021 | Number of cases in March, $V_j$ | Number of cases in April, $V_j$ | Number of cases in May, $V_j$ | Number of cases in June, $V_j$ | Number of cases in July, $V_j$ |
| --- | --- | --- | --- | --- | --- |
| 1 | 1357470 | 1711630 | 2083180 | 2206836 | 2236497 |
| 2 | 1364705 | 1731971 | 2085938 | 2209417 | 2237202 |
| 3 | 1374762 | 1745709 | 2088410 | 2211683 | 2237579 |
| 4 | 1384917 | 1755888 | 2090986 | 2213580 | 2237823 |
| 5 | 1394061 | 1769164 | 2097024 | 2214517 | 2238364 |
| 6 | 1401228 | 1784579 | 2105428 | 2215052 | 2238974 |
| 7 | 1406800 | 1803998 | 2114138 | 2216654 | 2239591 |
| 8 | 1410061 | 1823674 | 2119510 | 2218039 | 2240246 |
| 9 | 1416438 | 1841137 | 2122327 | 2219824 | 2240753 |
| 10 | 1425522 | 1853249 | 2124535 | 2221427 | 2241043 |
| 11 | 1438468 | 1861105 | 2129073 | 2222701 | 2241217 |
| 12 | 1451744 | 1872785 | 2135886 | 2223558 | 2241698 |
| 13 | 1460756 | 1887338 | 2143448 | 2223978 | 2242245 |
| 14 | 1467548 | 1903765 | 2150244 | 2224992 | 2242868 |
| 15 | 1477190 | 1921244 | 2153864 | 2226037 | 2243605 |
| 16 | 1489023 | 1936228 | 2156000 | 2227225 | 2244196 |
| 17 | 1504076 | 1946510 | 2160095 | 2228192 | 2244495 |
| 18 | 1519926 | 1953016 | 2165233 | 2229044 | 2244677 |
| 19 | 1535218 | 1961956 | 2170398 | 2229523 | 2245275 |
| 20 | 1546363 | 1974118 | 2175382 | 2229846 | - |
| 21 | 1554256 | 1990353 | 2179988 | 2230142 | - |
| 22 | 1565732 | 2004630 | 2182521 | 2230977 | - |
| 23 | 1579906 | 2017341 | 2183855 | 2231914 | - |
| 24 | 1596575 | 2025271 | 2186463 | 2232790 | - |
| 25 | 1614707 | 2030333 | 2189858 | 2233546 | - |
| 26 | 1632131 | 2038248 | 2193367 | 2233996 | - |
| 27 | 1644063 | 2047838 | 2196673 | 2234281 | - |
| 28 | 1652409 | 2059465 | 2199769 | 2234463 | - |
| 29 | 1662942 | 2069537 | 2201472 | 2235096 | - |
| 30 | 1674168 | 2078086 | 2202494 | 2235801 | - |
| 31 | 1691737 | - | 2204631 | - | - |

### Generalized SIR model and parameter identification procedure

The description of the generalized SIR-model and the exact solution of the set of non-linear differential equations relating the number of susceptible *S*, infectious *I* and removed persons *R* can be found in [14]. This solution uses the function

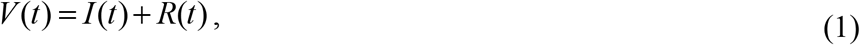

corresponding to the number of victims or the cumulative confirmed number of cases. Its derivative:

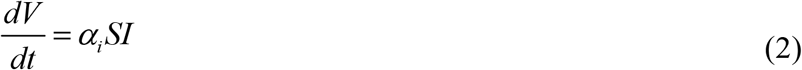

yields the estimation of the average daily number of new cases. When the registered number of victims *V*_*j*_ is a random realization of its theoretical dependence (1), the exact solution presented in [14] depends on five parameters (*α*_*i*_ is one of them). The details of the optimization procedure for their identification can be found in [15].

If we assume, that data set *V*_*j*_ is incomplete and there is a constant coefficient *β*_*i*_ ≥ 1, relating the registered and real number of cases during the *i-th* epidemic wave:

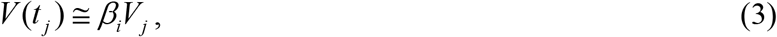

the number of unknown parameters increases by one. The procedure of their identification was presented in [24,25]. The values *V*_*j*_, corresponding to the moments of time *t*_*j*_ from the period March 11-24, 2021 have been used in [24, 25] to find the optimal values of these parameters corresponding to the tenth epidemic wave in Ukraine. In particular, the optimal value of the visibility coefficient *β*_10_ =3.7 was calculated.

### Monitoring changes in epidemic parameters and selection of epidemic waves

Changes in the epidemic conditions (in particular, the peculiarities of quarantine and its violation, situations with testing and isolation of patients, emergence of new strains of the pathogen) cause the changes in its dynamics, i.e. so known epidemic waves. To control these changes we can use daily or weekly numbers of new cases and their derivatives (see, e.g., [10, 12, 13]). Since these values are random, we need some smoothing (especially for daily amounts, which are also characterized by some weekly periodicity). For example, we can use the smoothed daily number of accumulated cases:

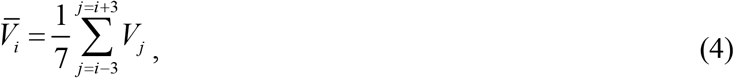

The first and second derivatives can be estimated with the use of following formulas:

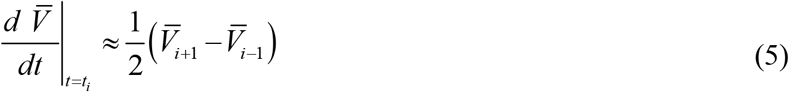

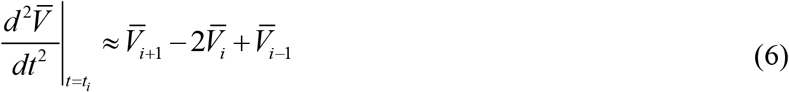

## Results and Discussion

First we have used the number of laboratory-confirmed cases (accumulated in the period the period *T*_*c11*_: May 23 – June 5, 2021 and presented in Table 1) for SIR simulations of the 11-th pandemic wave in Ukraine, i.e. we supposed that *β*_*i*_ =1 for *i*=11. The optimal values of parameters and other characteristics of this wave are calculated and listed in Table 2 (middle column). Comparison with the corresponding values for the tenth epidemic wave in Ukraine (see Table 2 in [25]) shows, that the optimal values of SIR parameters are very different for 11-th and 10-th pandemic waves. In particular, the estimations for the average time of spreading the infection 1/ *ρ*_11_ =4.1 days (in comparison with 1/ *ρ*_10_ =22.3 days for the tenth wave). The assessment of the 11-th epidemic wave duration (final day - August 25, 2021 corresponding the moment when the number of infectious persons becomes less that unit) is very optimistic (compare to March 29, 2022 for tenth wave).

**Table 2.** Visible and real characteristics of the eleventh COVID-19 pandemic wave in Ukraine. Results of calculations for optimal values of SIR parameters and other characteristics.

| **Characteristics** | 11<sup>th</sup> epidemic wave,<br>$i=11$,<br>$\beta_{11}=1$ | 11<sup>th</sup> epidemic wave,<br>$i=11$,<br>$\beta_{11}=20.376$ |
| --- | --- | --- |
| Time period taken for calculations $T_{ci}$ | May 23 – June 5, 2021 | May 23 – June 5, 2021 |
| $I_i$ | 10,190.8327995721 | 207,648.409124103 |
| $R_i$ | 2,180,167.16720043 | 44,423,086.1988759 |
| $N_i$ | 2,258,464 | 46018462.464 |
| $\nu_i$ | 60,891.3982283695 | 1,240,723.13030139 |
| $\alpha_i$ | 3.98540619731056e-06 | 1.95593158486001e-07 |
| $\rho_i$ | 0.242676955862249 | 0.242676955862287 |
| $1/\rho_i$ | 4.12070440082342 | 4.12070440082277 |
| $r_i$ | 0.996838194153353 | 0.996838194153390 |
| $S_{i\infty}$ | 31,667 | 645,251 |
| $V_{i\infty}$ | 2,226,797 | 45,373,211 |
| Final day of the epidemic wave | August 25, 2021 | September 20, 2021 |

The difference in saturation levels (final sizes) are not so large (*V*_11∞_ =2,226,797 and *V*_10∞_ =1,783,175). As of July 18, 2021 the registered number of COVID-19 cases in Ukraine (2,244,677) has already exceeded the saturation level of the eleventh epidemic wave. Since the difference recorded on day 43 after the last day of period used for the calculations *T*_*c11*_ is only 0.8%, we have another confirmation of the suitability of the generalized SIR-model for forecasting pandemic wave dynamics. The corresponding SIR curves (black lines), registered number of cases and the derivatives (5) and (6) (red markers) are shown in Fig. 1.

**Fig. 1.**
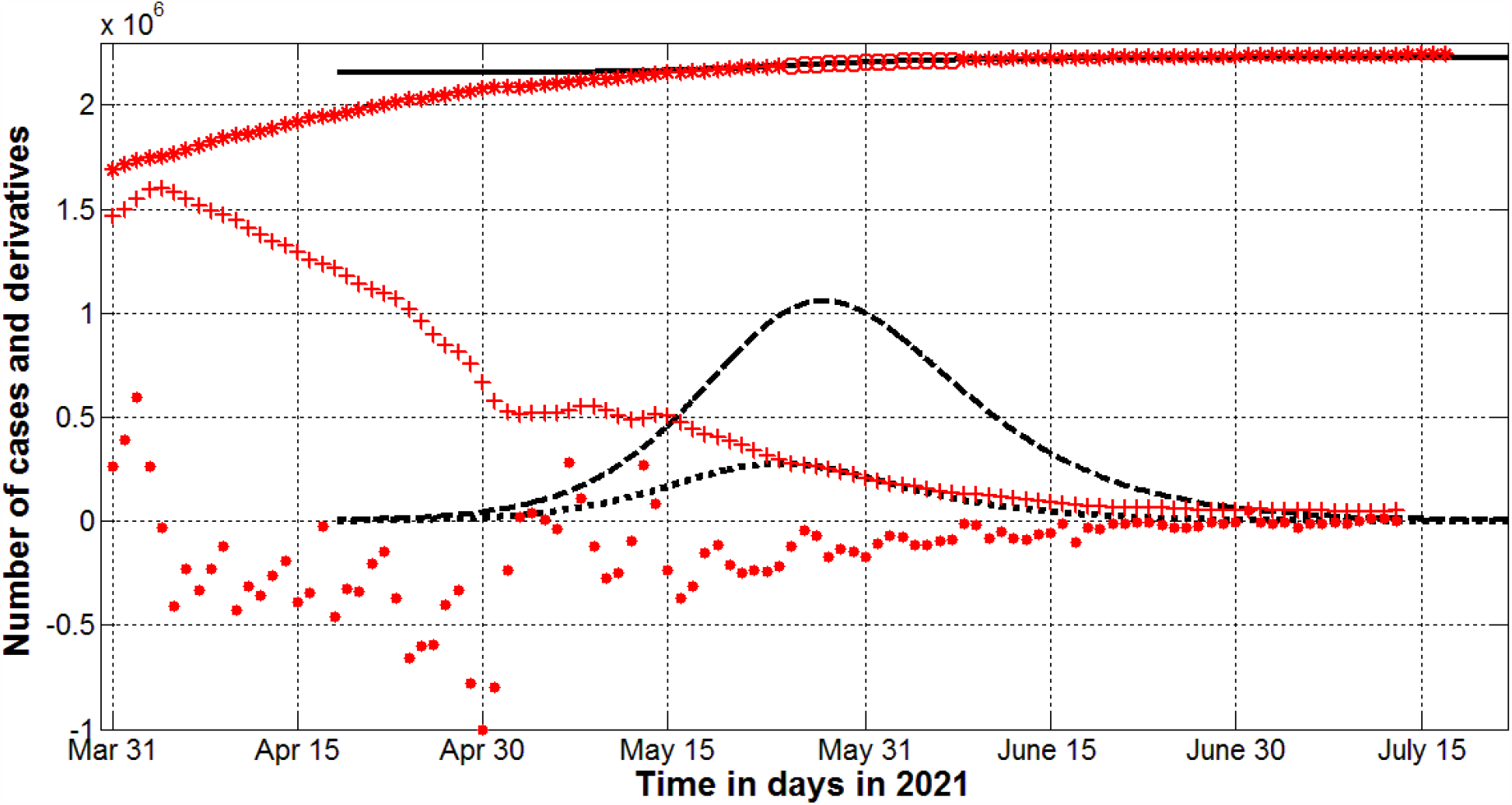
Visible COVID-19 epidemic dynamics in Ukraine in the spring and summer of 2021. The results of SIR simulations of the eleventh wave at *β*_11_ =1 are shown by black lines. Numbers of victims *V(t)=I(t)+R(t)* – solid lines; numbers of infected and spreading *I(t)* multiplied by 100 – dashed; derivatives *dV/dt* (eq. (2)) multiplied by 100 – dotted. Red “circles” and “stars” correspond to the accumulated numbers of cases registered during the period of time taken for SIR simulations (May 23 – June 5, 2021, Table 1) and beyond this time period, respectively. The red “crosses” and “dots” show the first derivative (5) multiplied by 100 and the second derivative (6) multiplied by 1000, respectively.

The jumps in values of the second derivative (6) shown in Fig. 1 by red “dots” illustrate the changes in the epidemic dynamics in the first half of May 2021 (probably connected with holidays). The values of the fist derivative (5) shown by red “crosses” are in good agreement with the theoretical estimation (2) (see black dotted line) for the period *T*_*c11*_ but deviate for the previous moments of time indicating significant changes in the epidemic dynamics. We can see also higher values of the first derivative (5) in comparison with estimation (2) (see black dotted line) after the period *T*_*c11*_. In particular, as of July 15, 2021 the estimation (5) yields value 13, but the average registered daily number of new cases (2) was 503. This fact and close to zero values of the second derivative (eq. (6), red “dots”) probably indicate the beginning of a new epidemic wave.

Last column of Table 2 illustrates the results of SIR simulations with the non-prescribed value of *β*_*i*_. The maximum of the correlation coefficient *r*_*11*_ was achieved at *β*_11_ =20.376. Thus, the vast majority of COVID-19 cases in Ukraine have become invisible (the real accumulated number cases is probably approximately 20 times higher than registered one). The real final size of the eleventh epidemic wave *V*_11∞_ is expected to be around 45.4 million and exceeds the population of Ukraine. If we multiply the number of registered cases (2,244,677 as of July 18, 2021) by the visibility coefficient *β*_11_ =20.376, the result 45.7 million is even higher. Probably, millions in Ukraine have already become re-infected and the nation achieved natural collective immunity (as of July 18, 2021 the percentage of fully vaccinated population was only 3.5%).

If the calculated value of the visibility coefficient *β*_11_ is correct, the mortality rate in Ukraine is not high. To estimate its value we can use the registered number of deaths (52,726 as of July 17, 2021). Probably, not all deaths caused by coronavirus have been reported, but in any case the visibility rate for them is lower than 20.376 due to the fact that critically ill patients usually go to hospitals and the corresponding deaths are properly recorded. Thus, the most optimistic estimation of the mortality rate is 52726*100%/(2244495*20.376)=0.12%.

As in the case of *β*_*i*_ =1, the optimal values of SIR model parameters are very different for tenth and eleventh epidemic waves (compare last columns in Table 2 and Table 3 from [25]). In particular, the differences are connected with the much higher value of the visibility coefficient (*β*_11_ =20.376 in comparison with *β*_10_ =3.7). The optimistic prediction for the 11-th epidemic wave ending (September 20, 2021) probably is not reliable since the beginning of a new epidemic wave is already visible in Fig.1). Ukrainians are also not guaranteed against the emergence or import of new coronavirus strains, which could cause new epidemic waves.

Optimal values of parameters presented in the last column of Table 2 allow us to calculate SIR curves corresponding to the real epidemic dynamics with the use of the exact solution presented in [14, 25]. The results are shown in Fig.2 by blue lines. The solid line represents complete accumulated number of cases (visible and invisible); the dashed line represents the complete number of infectious persons multiplied by 100, i.e. *I(t)x100*; dotted black lines represent the derivative *dV/dt* (which is an estimation of the real daily number of new cases) calculated with the use of (2) and multiplied by 100.

**Fig. 2.**
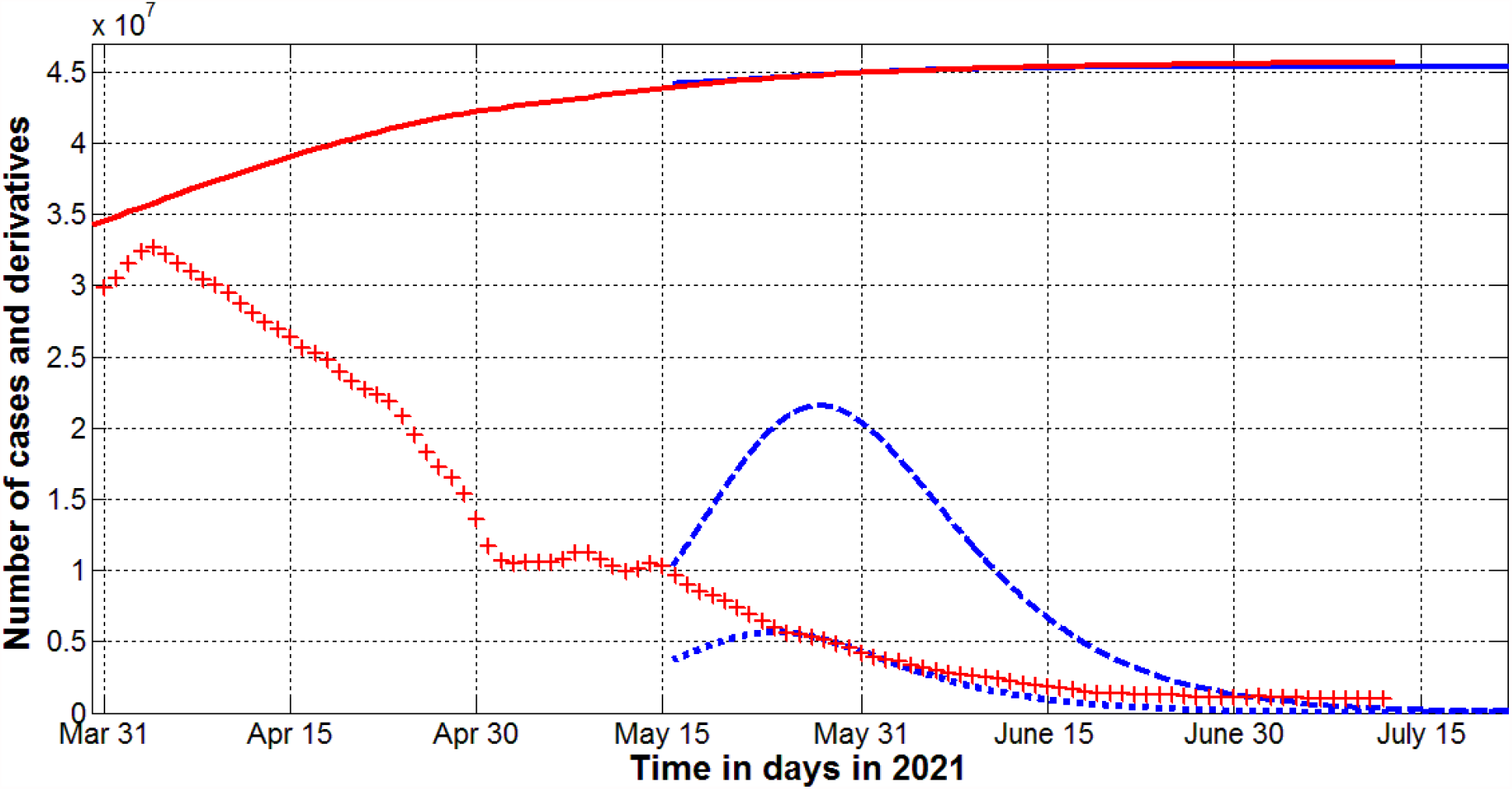
Real COVID-19 epidemic dynamics in Ukraine in the spring and summer of 2021. The results of SIR simulations of the eleventh wave at the optimal value *β*_11_ =20.376 are shown by blue lines. Numbers of victims *V(t)=I(t)+R(t)* – solid lines; numbers of infected and spreading *I(t)* multiplied by 100 – dashed; derivatives *dV/dt* (eq. (2)) multiplied by 100 – dotted. The red solid line shows smoothed accumulated number of laboratory-confirmed cases (eq. (4)) multiplied by the optimal value *β*_11_ =20.376. The red “crosses” show the first derivative (5) multiplied by 100*β*_11_.

Red “crosses” in Fig. 2 illustrate the estimation of the real average number of new daily cases for the eleventh wave, since the corresponding values were calculated by multiplying the derivative (5) by 100*β*_11_. These values are in good agreement with the theoretical estimation (2) for the period *T*_*c11*_ (May 23 – June 5, 2021) but deviate for the moments of time before and after this period (similar to the visible dynamics shown in Fig.1). The red line in Fig.2 represent the smoothed accumulated number of laboratory-confirmed cases (eq. (4)) multiplied by the optimal value of the visibility coefficient *β*_11_ =20.376 and is in very good agreement with the theoretical blue solid line.

The knowing of real sizes of the COVID-19 pandemic is very important to compare the effectiveness of vaccinations and a natural immunity. We can also estimate the probability of meeting an infected person with the use of simple formula, [10, 11]:

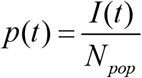

where *N*_*pop*_ is the volume of population. As of July 19, 2021 the theoretical estimations (with the use of parameters presented in the last column of Table 2) yield the value *I*=1656 (see the blue dashed line in Fig. 2). Then the probability *p* can be estimated as 0.00004. This value is much lower than corresponding estimation 0.015 for the end of March 2021, [25]. If the situation does not worsen, Ukrainians will be welcome guests in many countries even without vaccination.

## Data Availability

data is in the text

## Acknowledgements

The author is grateful to Oleksii Rodionov for his help in collecting and processing data.

